# Tuberculosis-Associated Respiratory Disability in Children, Adolescents, and Adults: Protocol for a Systematic Review and Individual Participant Data Meta-Analysis

**DOI:** 10.1101/2024.09.03.24313003

**Authors:** Silvia S. Chiang, Kamila Romanowski, James C. Johnston, Alex Petiquan, Mayara Bastos, Dick Menzies, Sierra Land, Andrea Benedetti, Faiz Ahmad Khan, Marieke M. van der Zalm, Jonathon R. Campbell

**Author notes:** **Corresponding author:** Jonathon R. Campbell, McGill University, Office 3D.55, 5252 Boulevard de Maisonneuve O, Montreal, Quebec, H4A 3S5. x.47454. These authors contributed equally (Silvia S. Chiang, Kamila Romanowski, Marieke M. van der Zalm, Jonathon R. Campbell).

## Abstract

**Background:** Approximately 2% of the global population has survived tuberculosis (TB). Increasing evidence indicates that a significant proportion of pulmonary TB survivors develop TB-associated respiratory disability, commonly referred to as post-TB lung disease (PLTD) and marked by impaired respiratory function, persistent symptoms, and activity limitations. However, the prevalence, risk factors, and progression of TB-associated respiratory disability throughout the life course are not well understood. To address these gaps, we will undertake a systematic review and individual participant-level data meta-analysis (IPD-MA) focusing on TB-associated respiratory disability in children, adolescents, and adults successfully treated for pulmonary TB.

**Methods and analysis:** We will systematically search MEDLINE, Embase, CENTRAL, Global Index Medicus, and medRxiv for original studies investigating TB-associated respiratory disability in people of all ages who have completed treatment for microbiologically confirmed or clinically diagnosed pulmonary TB. Authors of eligible studies will be invited to contribute de-identified data and form a collaborative group. Primary outcomes will be (1) abnormal lung function based on spirometry parameters and (2) chronic respiratory symptoms. We will estimate the overall and subgroup-specific prevalence of each outcome through IPD meta-analysis. Next, we will develop clinical prediction tools assessing the risk of future TB-associated respiratory disability at (i) the start of TB treatment and (ii) end of TB treatment for those without existing signs of disability. Finally, we will use stepwise hierarchical modelling to identify epidemiological determinants of respiratory disability.

**Ethics and dissemination:** This study has been approved by the ethics review boards at the Rhode Island Hospital (2138217-2) and the Research Institute of the McGill University Health Centre (2024-10345). Individual study authors will be required to obtain institutional approval prior to sharing data. Results will be disseminated through open-access, peer-reviewed publications and conference presentations.

**Prospero registration number:** CRD42024529906

**Strengths and limitations of this study:**

- An individual participant data meta-analysis allows for data harmonization to help overcome limitations of individual studies and aggregate meta-analysis, including small sample size, heterogeneity, and limited reporting of subgroups, such as age and other risk factors.
- We will be able to identify weaknesses in current reporting and recommend standards to support high-quality data collection and facilitate pooling of data.
- Key limitations include authors’ willingness to share data, representativeness of data contributed, and missing data.
- We will build an ongoing data collection platform to allow updating of evidence.
- Results will have implications for public health, clinical trial design, and clinical practice to support TB survivors.

## Introduction

Globally, over 10 million people develop tuberculosis (TB) each year,^1^ and as of 2020, an estimated 155 million individuals who had TB disease in the preceding 40 years were still alive.^2^ Approximately 86% of people who had TB disease had pulmonary TB,^1,3^ which even after successful treatment, may lead to respiratory sequelae, including restrictive lung disease, chronic obstructive pulmonary disease, and infectious pulmonary complications.^4,5^ This spectrum of TB-associated respiratory disorders, commonly known as post-TB lung disease (PTLD)—a misnomer as the pathogenesis occurs during TB disease—encompasses both respiratory impairment and functional limitations.^6,7^ Respiratory impairment is characterized by abnormal lung function, commonly measured through spirometry or structural assessments. Functional limitations relate to chronic respiratory symptoms and impact on daily activities and participation.^8^

A recent systematic review and aggregate-level meta-analysis, which included 61 original studies and over 40,000 participants, found 59% (95% confidence interval (CI) 49%, 69%) of people who survived TB had abnormal spirometry and 25% (95% CI 19%, 32%) reported breathlessness.^9^ Additional reviews have been conducted to identify risk factors for TB-associated respiratory disability, but consistent, robust associations have not been found.^10,11^ It remains unclear who is at the highest risk for TB-associated respiratory disability. Consequently, guidelines recommend screening all pulmonary TB survivors for TB-associated respiratory disability,^12–14^ a practice that is not feasible in the resource-limited settings where the majority of TB survivors reside.^1,2^ An improved understanding of risk factors and epidemiological determinants of TB-associated respiratory disability is crucial to enhancing the efficiency and feasibility of screening.

Another significant limitation in the existing evidence is the underrepresentation of children and adolescents in studies of TB-associated respiratory disability, along with a lack of age-disaggregated outcomes.^15^ Research across all age groups is essential due to the differences between children, adolescents, and adults in the host response to *Mycobacterium tuberculosis*, clinical presentation of pulmonary TB, and ongoing lung development in children and adolescents.^6^

Individual participant data meta-analysis (IPD-MA) can overcome many limitations of aggregate methods through harmonization of variables and outcomes, standardized analysis, analysis of subgroups not reported in primary studies such as age-stratified results, and increased sample size.^16^ Using IPD-MA, we can identify which subgroups are at the highest risk for TB-associated respiratory disability, enabling us to risk-stratify individuals for screening. Additionally, this approach allows us to explore how respiratory impairments and limitations evolve over time following treatment completion and to assess the underlying epidemiological determinants of TB-associated respiratory disability.

With a large body of literature now available, an IPD-MA is the ideal approach to improve the targeting and feasibility of screening strategies for TB-associated respiratory disability.

## Study aims and objectives

The overall aim of this systematic review and IPD-MA is to estimate the burden, risk, and determinants of TB-associated respiratory disability in children, adolescents, and adults treated for pulmonary TB. We will address the following specific objectives:

1. Estimate the prevalence of the two components of TB-associated respiratory disability: (i) impairment and (ii) symptoms and functional limitations. We will estimate prevalence among subgroups and examine temporal trends and risk factors.
2. Develop and validate risk prediction models to predict future TB-associated respiratory (i) impairment and (ii) symptoms and functional limitations.
3. Identify the epidemiological and clinical determinants of (i) respiratory impairment and (ii) respiratory symptoms and functional limitations, utilizing a conceptual framework and hierarchical approach.

## Methods and design

The systematic review and IPD-MA will be reported according to the 2020 PRISMA guidelines.^17^ This protocol has been reported according to the PRISMA-P guidelines^18^ (Appendix) and has been prospectively registered with PROSPERO (CRD42024529906). Any key changes or amendments will be documented there.

### Literature search and selection criteria

In preparation for this project, we conducted an initial literature search of MEDLINE, Embase, CENTRAL, Global Index Medicus, and medRxiv for original studies published between January 1st, 2004 and April 26th, 2024 using a comprehensive search strategy (Appendix) developed in collaboration with a medical librarian experienced in systematic reviews. We selected 2004 as the earliest publication year as there is minimal chance of individual participant-level study data being held for over 20 years. Two reviewers (KR and SL) screened titles, abstracts, full texts, and any studies identified as relevant from reviews and reference lists of eligible articles. Disagreements regarding inclusion or exclusion were resolved by a third reviewer (JRC). Retrieved references were uploaded into Zotero (Center for History and New Media, George Mason University), a reference management software, for de-duplication and then subsequently imported into Covidence (Veritas Health Innovation, Australia), a web-based platform designed to streamline the systematic review process. We will update this search through March 2025 to identify new studies published since our initial search.

Our study population will include children (0-9 years old), adolescents (10-19 years old), and adults (≥20 years old) with pulmonary TB that is microbiologically confirmed—through smear microscopy, mycobacterial culture, and/or molecular assays including GeneXpert—or clinically diagnosed, and in the case of children, meeting established clinical criteria.^19^ Wherever possible, our comparator population will include children, adolescents, or adults without a history of pulmonary TB. In the absence of a formal comparison group, the comparator will be assumed (e.g., for spirometry and other lung function measurements, international reference values for age, sex, and height will be used as the comparator).

Studies will be eligible for inclusion if they meet all of the following criteria: (1) prospective or retrospective cohorts, cross-sectional studies, or clinical trials, (2) included ≥10 participants who completed treatment for pulmonary TB, (3) measure and report at least one aspect of TB-associated respiratory disability, as detailed below, and (4) evaluate ≥80% of all participants for these outcomes to minimize selection bias associated with selective testing. We will include studies written in any language.

### Outcome measures

Our primary outcome will encompass the two aspects of TB-associated respiratory disability: (1) impairments to respiratory function and structure, and (2) chronic respiratory symptoms and/or limitations in daily activities and participation. These outcomes are based on the terminology used by the WHO International Classification of Functioning, Disability, and Health (ICF), a classification system that provides a standard language and framework for describing health and health-related states.^8^ According to the ICF, disability includes all negative aspects of health, including impairments in body function and structure and limitations in activities and participation.

Our primary outcome for impairments to respiratory function and structure will be abnormal lung function based on pre-bronchodilator spirometry parameters (**Table 1**). As secondary outcomes, we will also consider other impairments to respiratory function and structure, including (i) post-bronchodilator spirometry, (ii) pre- and post-bronchodilator oscillometry, (iii) measures of lung volume, (iv) diffusion capacity measurements, (v) other tidal breathing techniques, and (vi) presence of bronchiectasis.

**Table 1.** Outcome measures of tuberculosis-associated respiratory disability.

| Domain of disability | Outcome measure(s) |
| --- | --- |
| Impairments in respiratory function and structure | **Primary outcome** |
|  | Abnormal lung function, based on pre-bronchodilator spirometry parameters, defined as FEV1, FVC, and FEV1/FVC and classified based on specific disease patterns, including obstructive, restrictive, or mixed pattern. |
|  | **Secondary outcome(s)** |
|  | (i) post-bronchodilator spirometry, (ii) pre- and post-bronchodilator oscillometry, (iii) measures of lung volume, (iv) diffusion capacity measurements, (v) other tidal breathing techniques, or (vi) presence of bronchiectasis. |
| Symptoms and/or limitations in daily activities and participation | **Primary outcome** |
| | Chronic respiratory symptoms, defined as experiencing at least one of the following symptoms $\geq 2$ days per week: cough, sputum production, wheeze, dyspnea, and/or chest pain, measured using symptom assessment or validated questionnaire (e.g., SGRQ) |
|  | **Secondary outcome(s)** |
|  | (i) Grades 3-5 on MRC Dyspnoea Scales, (ii) grades 2-4 on the modified MRC Dyspnoea Scale, (iii) values higher than 'moderate breathlessness' on Borg Dyspnoea Scale; or (iv) 6MWT result below predicted lower limit of normal, calculated using standard equations based on age, sex, height, and weight. |
| Abbreviations: 6MWT, 6-minute walk test; FEV1, forced expiratory volume in 1 second; FVC, functional vital capacity; MRC, Medical Research Council; SGRQ, St. George's Respiratory Questionnaire. |  |

Our primary outcome for symptoms and/or limitations in daily activities and participation will be chronic respiratory symptoms (**Table 1**). As secondary outcomes, we will also consider: (i) grades 3-5 on the MRC Dyspnoea Scales,^20^ (ii) grades 2-4 on the modified MRC Dyspnoea Scale,^21^ (iii) values higher than ‘moderate breathlessness’ on the Borg Dyspnoea Scale;^22^ and (iv) 6MWT result below predicted lower limit of normal, calculated using standard equations based on age, sex, height, and weight.^23^

### Invitations to authors of eligible studies

Corresponding authors of eligible studies will be contacted via email and invited to join the collaborative group and share their de-identified study data. If there is no response from the author within four weeks, we will try a second time. If authors do not respond or indicate that the data are unavailable or cannot be shared due to access restrictions, we will note that data are unavailable.

After accepting the invitation to collaborate, signing a data transfer agreement, and obtaining institutional approvals, authors will transfer their data securely via the Nextcloud Hub (Nextcloud GmbH, Germany). Data will be housed on a secure server at McGill University.

### Data collection, processing, and management

From each eligible study, we will collect study-level and individual-level variables. Study-level variables will include country, funding source, country-level health characteristics, study design, population, aims, recruitment period, and test(s) used to measure outcomes. Individual-level variables will include demographic, clinical, radiographic, microbiologic, and outcome data (**Table 2**). All received study data will be reviewed for missing, incomplete, or implausible data and compared against published information; authors will be further consulted for clarifications. Prior to processing, we will exclude all participants with missing information on age or those without an outcome measure of interest. We will standardize outcomes and covariates between studies using systematic harmonization methodology.^24^ Study-specific data items will be processed into a common format for analysis.

**Table 2.** Individual-level variables to be requested from study authors.

| Category | Variable | Proposed stratification, if relevant |
| --- | --- | --- |
| Demographic | Age (years) at TB treatment initiation | <5, 5-9, 10-14, 15-19, 20-35, 36-54, 55-75, >75 |
|  | Sex | Male, Female |
|  | Gender | Male, Female, Other |
|  | Height | For children, we will consider various measures for height and weight, such as weight-for-age or height-for-age z-scores |
|  | Weight |  |
|  | BMI (kg/m<sup>2</sup>) | <18.5, 18.5-24.9, 25-29.9, >30 |
|  | Active smoking exposure<sup>^</sup> | Never, Former, Every day, Someday<sup>^</sup> |
|  | Smoking duration (years), *for those who are every day, someday, and former smokers* | <1, 1-5, 6-10, 11-19, >20 |
|  | Antenatal passive smoking exposure<sup>#</sup> | Yes, No |
|  | Postnatal passive smoking exposure<sup>#</sup> | Yes, No |
|  | Environmental biomass exposure, *defined as cooking or heating living areas with solid fuel* | Yes, No |
|  | Adult education level | Less than high-school, High school, University, Post-Graduate |
|  | Area of residence | Urban, Rural |
| Clinical, biological, radiographic, and treatment | HIV status | HIV on ART, HIV without ART, HIV-negative |
|  | CD4 count, *for those living with HIV* | <350, +350 |
|  | Pre-existing asthma | Yes, No |
|  | Pre-existing COPD | Yes, No |
|  | Pre-existing other respiratory comorbidities | Yes, No |
|  | Diabetes | Yes, No |
|  | Previous TB treatment episodes | Yes, No |
|  | Treatment delay | Yes, No |
|  | Diagnostic certainty of TB diagnosis | Clinical diagnosis, Microbiological diagnosis |
|  | Drug resistance pattern of *M. tuberculosis* | Drug-susceptible, Isoniazid-monoresistant, Rifampicin/multidrug resistant, Extensively drug-resistant |
|  | TB disease severity at diagnosis<sup>*</sup> | Severe disease, Non-severe disease |
|  | TB treatment duration | <7 months, +7 months |
| Outcome measures | All outcome measures described in **Table 1** |  |
|  | Months between TB treatment end and outcome measure | During treatment, <6, 6 -12, 12-24, >24 |

<sup>#</sup>Obtained for paediatric and adolescent populations
<sup>^</sup>Every day smoker defined as an adult who has smoked 100 cigarettes in his or her lifetime and who currently smokes cigarettes, while someday smoker defined as an adult who has smoked at least 100 cigarettes in his or her lifetime, who smokes now, but does not smoke every day, as defined by the CDC National Centre for Health Statistics.
<sup>\*</sup>For children, TB disease severity will be assessed according to WHO criteria, based on bacteriological burden and/or radiographic findings. Non-severe pulmonary disease in children will be defined as intrathoracic lymph node TB without airway obstruction; uncomplicated TB pleural effusion or paucibacillary, non-cavitary disease confined to one lobe of the lungs and without a miliary pattern. For adults, those acid fast bacilli smear positive or with cavitation on chest x-ray will be considered to have severe disease.
Abbreviations: BMI, body mass index; HIV, human immunodeficiency virus; ART, antiretroviral treatment; COPD, chronic obstructive pulmonary disease

We will quantify individual-level missing data in each study. We will impute missing data using multiple imputations while respecting participant clustering by source study.^25^ We will generate 20 imputed datasets, each undergoing 25 between-imputation iterations. All analyses will be done in each of the 20 imputed datasets, with estimates pooled according to Rubin’s Rules.^26^ No imputations will be done for outcome measures.

### Risk of bias assessment

Two study investigators will independently assess the risk of bias in included studies using an adapted version of the ROBINS-E tool (**Table 3**).^27^ We will evaluate the following criteria at the study level: (1) selection of participants, (2) measurement of exposure, (3) confounding, (4) post-exposure interventions, (5) measurement of outcome, and (6) missing data. Any disagreements will be resolved through discussion or consultation with a third study investigator. For each subdomain, we will assign a risk of bias of low, medium, high, very high, or uncertain. As no detailed guidance has been developed on providing an overall risk of bias for each study, we will not give an overall risk of bias but rather discuss the potential impacts of identified sources of biases on the interpretation of our findings.

**Table 3.** Risk of bias criteria.

| **Domain** | **Question(s)** |
| --- | --- |
| **Risk of bias due to confounding**<br>*Key confounders: environmental exposures, smoking exposure, age, weight, sex, HIV status, and pre-existing respiratory comorbidities.* | Did the author(s) consider all important confounders in the design of the study (i.e., matching)?<br>Did the author(s) consider all important confounders in the analysis (i.e. stratified analyses, multivariate models)?<br>Did the author(s) collect information on all important confounders?<br>Were confounding factors measured validly and reliably (i.e., secure record)? |
| **Risk of bias arising from measurement of the exposure** | Was TB diagnosis based on standardized definitions for diagnostic classification?<br>Were results presented stratified by microbiological confirmation and clinical diagnosis?<br>Was TB treatment data ascertained validly and reliably (i.e., secure record)? |
| **Risk of bias in selection of participants into the study** | Was the TB-exposed cohort selected randomly or using a consecutive sample?<br>Was the comparator population selected from the same community as the TB population? |
| **Risk of bias due to post-exposure interventions**<br>*Post exposure interventions: Pulmonary rehab, steroids, surgery* | Did the study report if participants received any post-exposure interventions that may have decreased the sensitivity of the outcome?<br>Did the analyses attempt to correct for the effect of any post-exposure interventions? |
| **Risk of bias due to missing data** | Were complete data on exposure status available for all, or nearly all (>90%), participants?<br>Were complete data on outcome status available for all, or nearly all (>90%), participants?<br>Were complete data on collected confounders available for all, or nearly all (>90%), participants? |
| **Risk of bias arising from measurement of the outcome** | Was TB-associated respiratory disability measured using standardized methods (i.e., validated tools, according to guidelines)?<br>Did the timing of the outcome vary significantly among participants? |

Abbreviations: HIV, human immunodeficiency virus; TB, tuberculosis

### Grading the strength of existing evidence

We will assess the existing evidence using categories in the Cochrane Handbook, which considers inconsistency, indirectness, imprecision, and bias.^28^ Bias will be assessed for each individual study using the risk of bias assessment described above, and globally we will assess publication bias visually using Egger plots and forest plots for our two primary outcomes. Inconsistency will be assessed according to the similarity in the magnitude and direction of effects across studies; we will use stratified analyses to evaluate potential sources of heterogeneity. Indirectness will be evaluated based on applicability (tests or evaluations performed, time frame) and participant selection.

Imprecision will be measured according to precision/confidence intervals around estimates, while considering sources of heterogeneity. Based on the above considerations, the strength of the existing evidence will be graded as high (no concerns with any of the above considerations), moderate, low, or very low (based on the number of concerns). In line with Cochrane, we may adjust certainty based on other factors, such as large effects, observing a dose-response, or presence of plausible confounding.^28^

### Statistical analysis

Unless otherwise specified, we will conduct analyses on the imputed datasets and complete case analysis. Stratified analyses will be done on the variables included in **Table 2**. When conducting stratified analyses, we will stratify only on known covariate information (e.g., for analyses stratified on female sex, only participants reported to be female will be included; those with imputed sex will be excluded). All analyses will be done using R (The R Project for Statistical Computing, the latest version available at the time of analysis start).

#### Objective 1

We will calculate the overall and subgroup-specific prevalence of TB-associated respiratory disability in two stages.^29^ In the first stage, we will use the individual participant data to estimate the proportion of people with respiratory disability and the standard error within each individual study. In the second stage, we will pool the logit-transformed proportions of disability across studies using generalized linear mixed models. We will back-transform the pooled estimates and standard errors to obtain prevalence estimates and 95% CIs and generate forest plots to compare prevalence estimates across strata.^30,31^ We will use I^2^,τ^2^, and prediction intervals to describe variability resulting from between-study heterogeneity.

Next, we will use one-stage IPD-MA to estimate the prevalence ratio for the outcomes of (i) respiratory impairment and (ii) symptoms and activity limitations across strata. We will use generalized linear mixed log-binomial models, adjusted for variables in **Table 2**, plus the time point of outcome measurement to see how outcomes change over time. We will account for clustering at the study and participant level. Heterogeneity will be assessed with I^2^.^32^

Based on findings from a previous aggregate meta-analysis, we estimated the sample size required to estimate a prevalence of 59% for respiratory impairment and 25% for chronic symptoms and/or activity limitations, with an estimated average cluster size of 140.^9^ Under the assumption that the intraclass correlation coefficient between studies was 0.03,^33,34^ with a power of 80%, type I error rate of 5%, and absolute precision of 10%, we will require a sample size of 481 for our outcome of respiratory impairment and 373 for our outcome of symptoms and limitations. This suggests we will likely have sufficient power for our primary outcomes and for several key subgroups.

#### Objective 2

We will develop risk prediction models adhering to TRIPOD (Transparent Reporting of a Multivariate Model for Individual Prognosis or Diagnosis)^35^ guidance to predict the future risk of TB-associated respiratory disability at (i) the start of TB treatment, (ii) the end of TB treatment among those who do not already have evidence of TB-associated respiratory disability, and (iii) the end of TB treatment among those with evidence of TB-associated respiratory disability. The latter two outcomes are important as existing data suggest up to one-quarter of people who go on to develop TB-associated respiratory disability are asymptomatic at the end of treatment,^36^ and it is of interest to determine if there are people at higher risk of transient disability.

For our prediction models, we will use a sequential method to select candidate predictors. First, we will use *a priori* selection, based on consensus among investigators, published data, and expert opinion, to sequentially include the chosen predictors in a layered fashion.^37^ Next, we will use elastic net penalized regression models to identify key predictors from amongst the larger set of variables available to evaluate if our *a priori* predictors were missing highly predictive variables.^38^ We will evaluate predictive performance using discrimination and calibration measures and validate the models using the internal-external cross-validation (IECV) framework.^39^

To estimate the required sample size for risk prediction models, we used a conservative predictive performance estimate of a previous model (0.71)^40^ and estimated sample size with 20 candidate predictors. If the proportion of survivors with symptoms and/or activity limitations is 25%, we require a sample size of 1836 to develop a prediction model; if the prevalence of respiratory impairment is 59%, we require a sample size of 1425.^41^

#### Objective 3

To improve our understanding of the contribution of TB to respiratory disability, as opposed to other sociodemographic, clinical, and behavioural determinants, this specific aim will include only studies with a comparator group of people who never had TB disease. We will use a stepwise hierarchical modelling approach, which can systematically delineate confounding and mediating factors from our outcome of respiratory disability.^42^ Our modelling approach will follow the conceptual framework in Figure 1, with factors organized according to how proximate each factor is to our outcome of respiratory disability.

**Figure 1.**
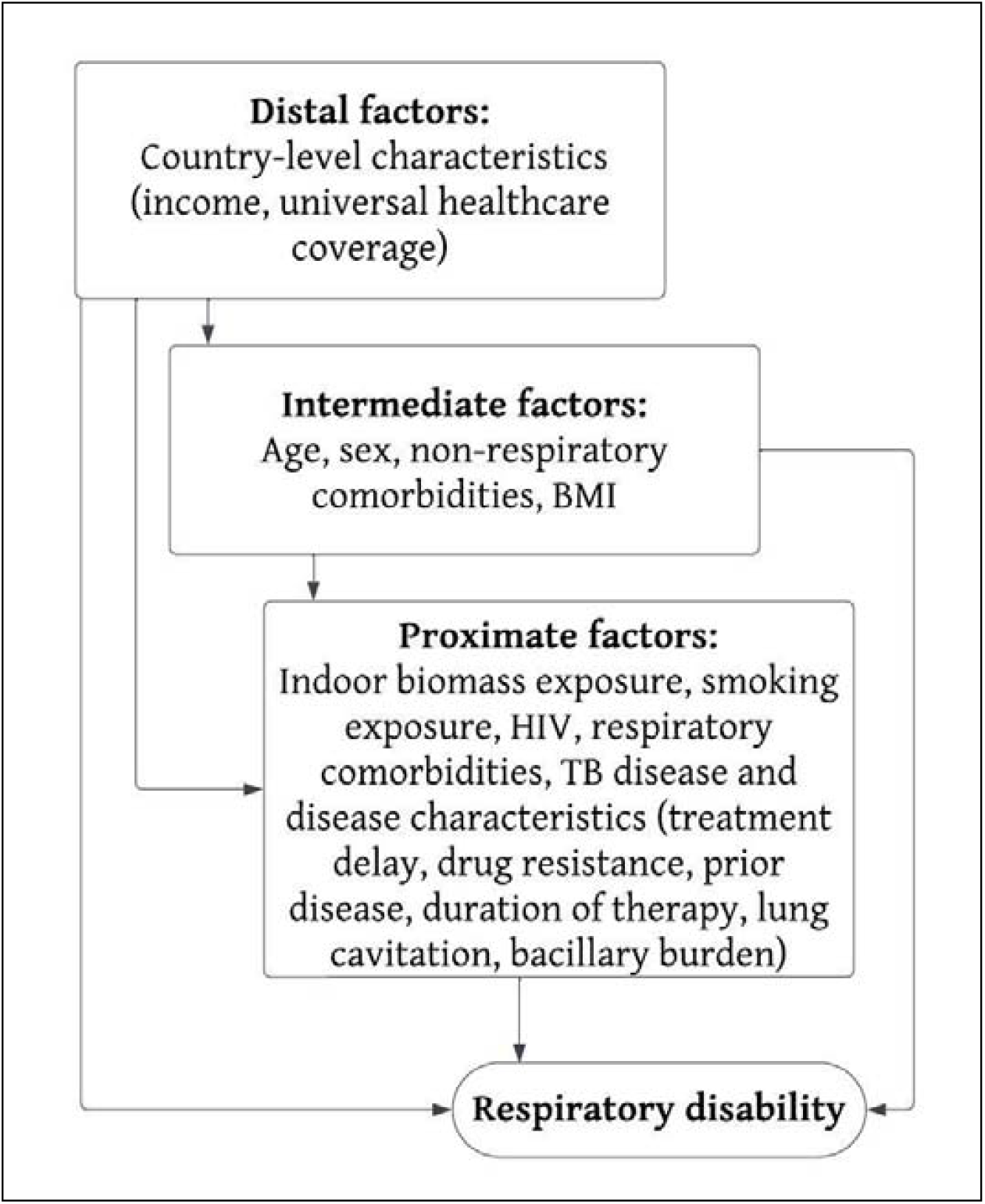
Epidemiological determinants of tuberculosis-associated respiratory disability Abbreviations: HIV, human immunodeficiency virus; TB, tuberculosis; BMI, body mass index.

We will use generalized linear log-binomial models with random effects for each study. Models will be structured by first including distal determinants and subsequently including intermediate and proximate determinants in a hierarchical fashion. In this framework, the impact of TB on respiratory disability is mediated by disease-related factors such as bacillary burden and length of treatment. We will ultimately estimate the effect of TB on the outcomes of (i) respiratory impairment and (ii) symptoms and/or limitations in activity adjusted for confounders and not mediated through proximate determinants.

### Patient and public involvement

No direct patient or public involvement has taken place during the development of this protocol. However, we will work with the Community Advisory Boards of the McGill TB Centre (Canada) and the Desmond Tutu TB Centre (South Africa), as well as TB survivor networks and advocacy groups such as STOP TB USA, STOP TB Canada, TB Proof, and the Child and Adolescent Working Group of the WHO/Stop TB Partnership during the interpretation and dissemination of the study results.

### Governance

All contributing authors will sign data sharing agreements with the institution hosting the secure data server (The Research Institute of the McGill University Health Centre), which outlines standards of data security, management, and ownership (Appendix 2). The initial length of this data-sharing agreement will be five years. All data will be treated as confidential and will remain the property of the contributing institution; data can be withdrawn at any time. All contributing authors will enter a consortium, which collaboratively completes the outlined analyses. JRC will act as the data custodian and will be the primary point of contact between consortium members.

An oversight committee, consisting of 7 members, will be established with the data custodian (JRC) acting as additional member in a non-voting role. For the first four years, this committee will comprise the other three study principal investigators (SSC, JCJ, MMvdZ), and four elected members of the consortium; thereafter, all oversight committee members will be elected. As the data custodian, JRC will remain on the oversight committee in an unelected position with a non-voting role. The oversight committee has three specific roles: (i) review requests for individuals or organizations to have access to the IPD for the purposes of agreed-upon analyses; (ii) review and discuss proposals by members of the consortium for projects or analyses not outlined in this protocol; and (iii) review opportunities and requests for authorship and/or participation in analyses to ensure contribution and opportunity is equitable among the consortium. Issues deemed significant by the oversight committee may be brought forth to all consortium members for input.

### Ethical considerations

The IPD-MA was approved by institutional ethics review boards at Rhode Island Hospital, U.S.A. (2138217-2) and The Research Institute of the McGill University Health Centre, Canada (2024-10345). Individual studies will share de-identified data and follow institutional and national guidelines for data sharing.

## Discussion and conclusions

Given that approximately 1 in 50 people globally have survived pulmonary TB and the growing demand for evidence to inform post-TB care,^2^ this IPD-MA has the potential to significantly impact recommendations for TB-associated respiratory disability. Through an IPD-MA, we will provide evidence to help identify who will be most at risk for TB-associated respiratory disability. Our analyses to risk stratify individuals will be essential to inform efficient recruitment into randomized clinical trials of TB-associated respiratory disability-related interventions and can support patient selection for clinical evaluation and follow-up after successful TB treatment in resource-limited settings. Given the diversity of factors that may cause the spectrum of signs and symptoms that comprise TB-associated respiratory disability, our analysis of epidemiological determinants will help elucidate key aspects beyond TB for which we might intervene.

Our IPD-MA does have limitations and challenges. A major challenge is mapping and harmonizing variables due to inconsistencies in data collection methods, variable definitions, and missing information across studies. We will partner with Maelstrom Research, a group with recognized expertise in data harmonization, for rigorous data pre-processing and harmonization techniques.^24^ Given the heterogeneity in data collection across studies in all fields, our work on data harmonization will also help us make recommendations to improve the uniformity, accuracy, and completeness of data collection for future studies. These will further facilitate future collaborative research, including updates of this project.

Authors’ willingness to share data is another barrier, and data for certain regions or subgroups may be challenging to obtain.^43^ We have budgeted resources to facilitate data sharing for authors from all settings and have established guidelines in our data-sharing agreements to promote transparency and collaboration. We will also establish a data repository, allowing us to include more data as it becomes available. Authors contributing data likely come from large centres, with adequate resources for monitoring and evaluation post-treatment. This may not be generalizable to lower-resource settings; however, we have designed our risk stratification analysis to provide evidence to programs of all resource levels. Bias related to participant selection, data collection, or reporting may affect the validity of our analyses. We are using a robust bias assessment tool to structure our bias and study quality assessment and have implemented inclusion criteria to minimize participant selection. Finally, pediatric subgroup analyses may be limited by small sample size. The inclusion of children and adolescents in this IPD-MA supports advocacy efforts for more data in this group and allows for the inclusion of currently underway studies.

In summary, we will conduct an IPD-MA of TB-associated respiratory disability among TB survivors of all ages to determine who is most at risk, help predict those who might benefit from screening, and improve our understanding of TB’s contribution to respiratory disability. The insights gained from these analyses may enhance strategies for detecting and preventing TB-associated respiratory disability and inform the design of clinical trials of interventions to prevent TB-associated respiratory disability.

## Data Availability

All data produced in the present study are available upon reasonable request to the authors

## Declarations

### Ethics approval and consent to participate

This study has been approved by the ethics review boards at the Rhode Island Hospital (2138217-2) and the Research Institute of the McGill University Health Centre (2024-10345). Individual study authors will be required to obtain institutional approval prior to sharing data.

### Consent for publication

NA

### Availability of data and materials

Once the objectives outlined in this Protocol are achieved, individuals or groups from within or outside the TB-Residual consortium may contact the Project Leads to propose further studies.

## Authors’ contributions

Conception and Design: SSC, KR, JCJ, MMvdZ, JRC

Intellectual Contribution and Interpretation: All authors

First Draft: SSC, MMvdZ, KR, JRC

Revisions: All authors.

All authors read and approved the final manuscript.

## Appendix

**Appendix Table 1.**
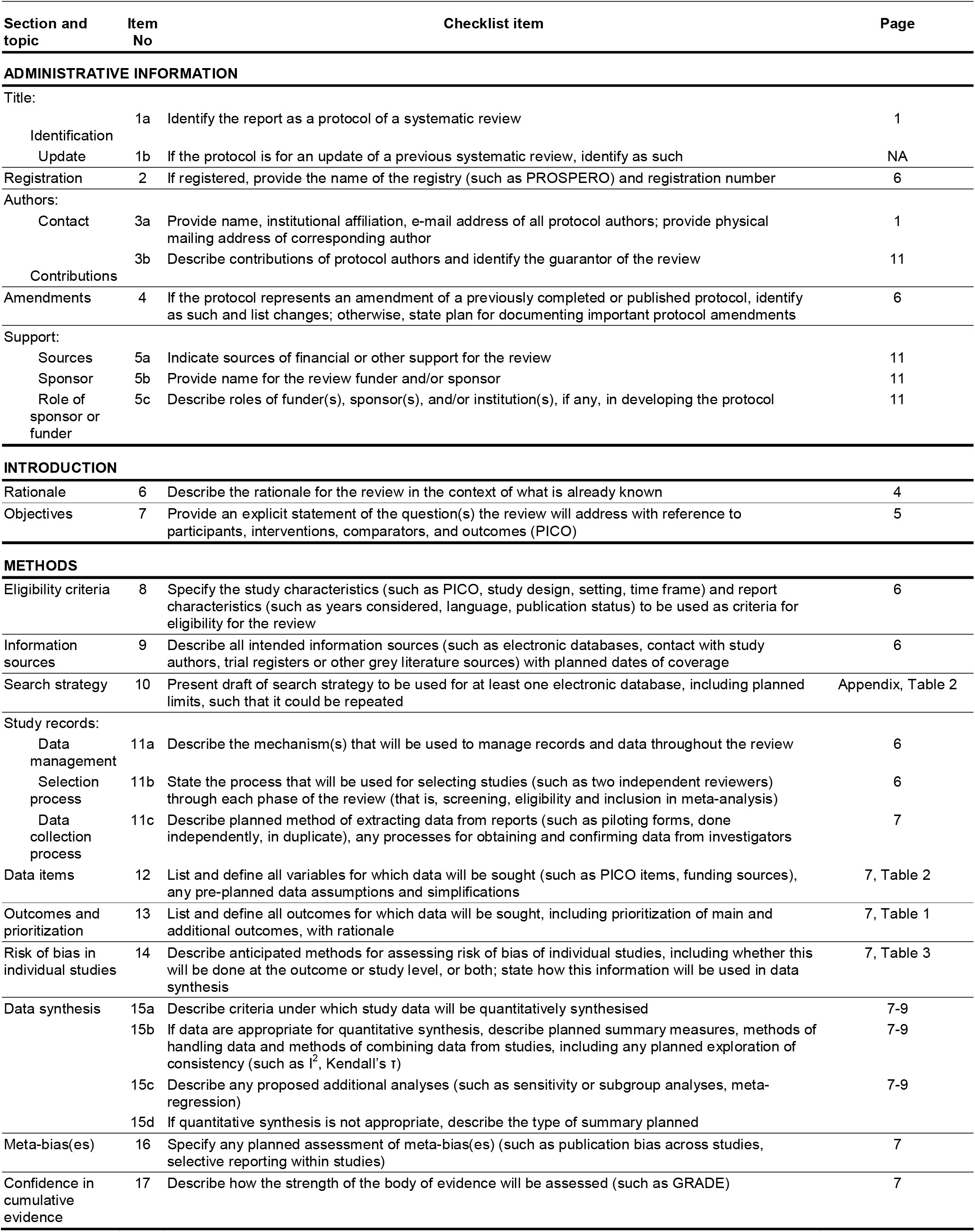
PRISMA-P (Preferred Reporting Items for Systematic review and Meta-Analysis Protocols) 2015 checklist: recommended items to address in a systematic review protocol*.

**Appendix Table 2.**
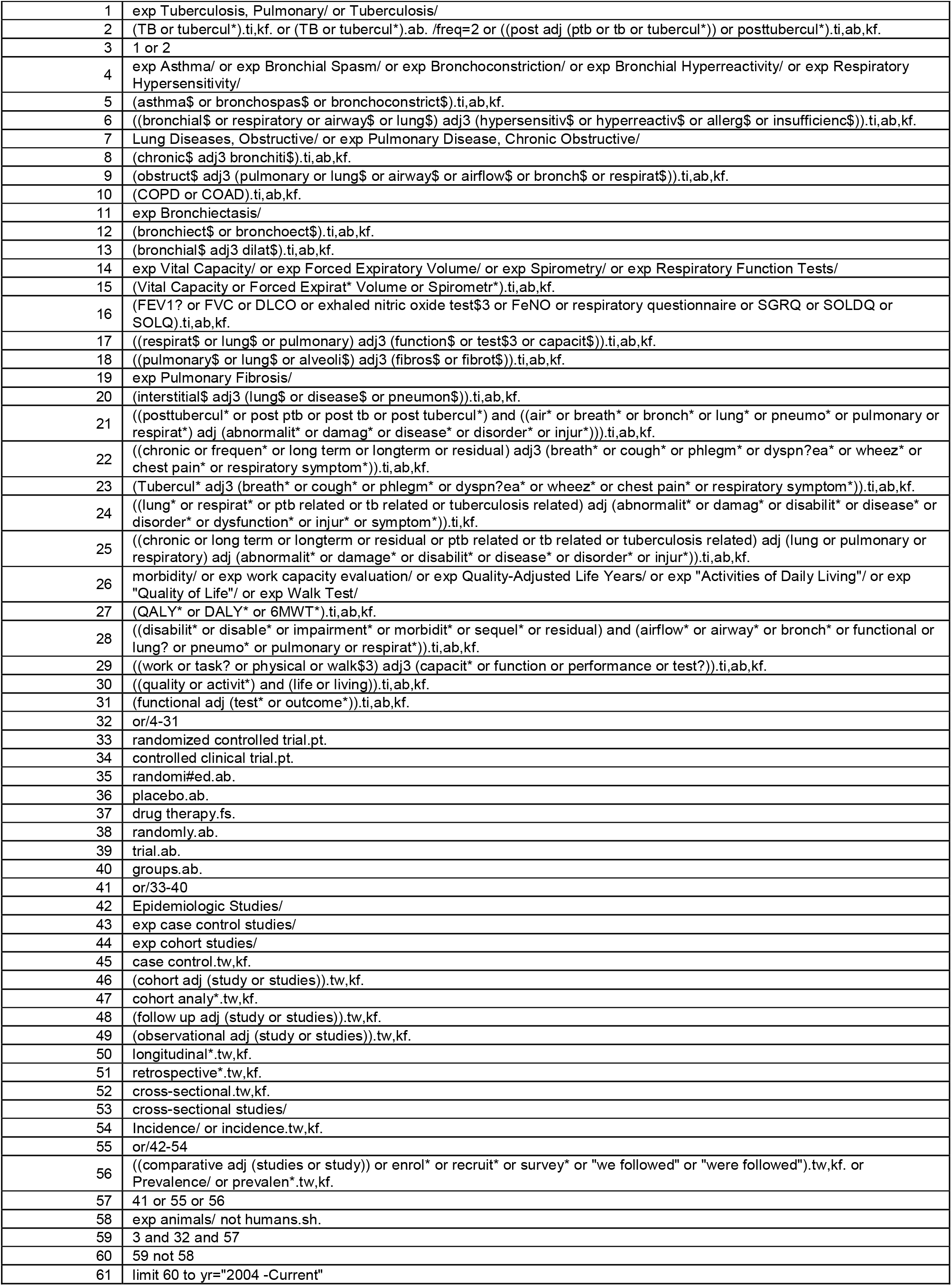
Search strategy for MEDLINE ALL (Ovid)

