## Appendix 2 for "Tuberculosis-Associated Respiratory Disability in Children, Adolescents, and Adults: Protocol for a Systematic Review and Individual Participant Data Meta-Analysis"

DATA SHARING AGREEMENT (DRAFT)

**LETTER OF AGREEMENT FOR A SHARED INDIVIDUAL PARTICIPANT DATA PLATFORM FOR POST-TUBERCULOSIS CONSEQUENCES (POST-TB-IPD)**

This letter of agreement is between Dr. Jonathon Campbell for the McGill University group who will act as Data custodian (hereafter referred to as the “McGill Group”) for ***The shared individual participant data platform for post-tuberculosis consequences (POST-TB-IPD) – present and future*** and the Institution or Group contributing data **** INSERT NAME OF INVESTIGATOR AND INSTITUTION *** / Data contributor (hereafter referred to as the “Data Contributor”), regarding the transfer and use of data collected by the investigator.

The McGill Group and the Data Contributor agree to collaborate on the ***POST-TB-IPD*** according to the terms in this letter and those set out in the full project protocol, which is attached as Annex 2.

The initial duration of this agreement is for a period of five years (<potentially updated based on discussion>). The agreement can be renewed for a mutually agreed upon duration at the end of this initial period. The data contributor may rescind this agreement at any point.

*The McGill Group agrees to respect the principles of the POST-TB-IPD:*

1. Obtain approval from the Research Ethics Board of the Research Institute of the McGill University Health Centre for the maintenance of this database (MUHC-REB-2024-10345, Approved June 7, 2024).
2. Receive data only through secure means, mutually agreed upon by the McGill Group and Data Contributor to be NextCloud (<edit based on agreed platform>).
3. Respect the confidentiality of all data received. The McGill Group will not attempt to identify participants, nor contact participants directly. The data provided will be non-nominal and no identification of the individual participant’s identity can be made at any time. This includes ensuring that, in the analysis, particularly of small groups (<5 participants), individual identities cannot be inferred from presentation of the results. The Institutions or Groups contributing data will be apparent in the data set to anyone performing analysis, through a contributor-level identifier attached to all participant data contributed, and the identity of institutions/groups may be revealed in descriptive analyses. However, analytic results such as correlates of post-tuberculosis consequences, or specific outcomes (e.g. abnormal spirometry) will not be reported at the level of specific countries, institutions, or groups contributing data.
4. The data contributed continues to be owned by the individual, group, country or institution that has contributed the data. This means the data can be withdrawn from the POST-TB-IPD at any time. The McGill Group will destroy received data at such a time.
5. Data and analysis will be accurate and of the highest possible quality. The individual, group, or institution contributing data remains responsible for the accuracy of the data submitted. The Data Custodian will take all measures to ensure that the quality is improved through coherency checks and other quality assurance measures in place to safeguard the wholesomeness of the data.
6. Data will be kept by the Data Custodian in a secure environment so that it cannot be accessed except by the Data Custodian, and approved co-investigators and analysts (Dr. Silvia Chiang, Dr. Marieke Van der Zalm, Dr. James Johnston, and Dr. Kamila Romanowski). Additional individuals may be added to the list of individuals able to access contributed data contingent on approval of the POST-TB-IPD Oversight Committee (See Annex 1 for details of the membership, selection process and responsibilities of this POST-TB-IPD oversight committee).
7. The data housed by the McGill Group as part of the POST-TB-IPD are to be shared for use, under certain conditions and after approval by the POST-TB-IPD Oversight Committee. This means that, after a process of review and approval, the data can be accessed for analysis by contributing members of the shared POST-TB-IPD, as well as other not-for-profit organizations or institutions, or national TB programs.
8. All data contributors agree that for the specific purpose of WHO guidelines development—a global public health good—use of the data base will not require review by the Oversight Committee or individual data contributors. Instead, the statistical analysis plans, including specific questions or objectives will be determined by the Guidelines Development Groups (GDG) convened by WHO for guideline development. Hence for all analyses to inform WHO guidelines, the GDG’s of WHO will have oversight and approve use of the shared POST-TB-IPD.
9. All proposed data analyses will be reviewed by the Data Custodian, to advise on feasibility and data completeness and quality issues related to the statistical analysis plan (SAP).
10. The McGill Group may also perform data analysis, but only after this use has been approved by the POST-TB-IPD Oversight Committee, or by a GDG of WHO (i.e. the McGill group must follow the same procedures as any other group or researcher).
11. The Research Ethics Board of the Research Institute of the McGill University Health Center that oversees the institution that is serving as the Data Custodian will also review all uses of data not covered under initial ethical approvals.
12. Access to the data for all other uses, and for any analyses that will lead to eventual publication (other than publication of WHO guidelines) will be regulated by the POST-TB-IPD Oversight Committee comprised of representatives of data contributors and the Data Custodian.
13. The data shall not be used for profit, nor sold to any third party.
14. If access to the secure environment housing the data is provided to an individual, group or institution, they must not remove data from the secure environment or use it for any unauthorized purpose.
15. Queries about the data from outside organizations performing data analysis will be sent to the Data Custodian, who may choose to refer the questions to the original data contributors. There will not be any direct queries between data analysts and data contributors. This will ensure that any additions or corrections to the data are well documented, and that the POST-TB-IPD database is as accurate and up-to-date as possible, and that at any time, there is only one version of the database.

*The Institution or Group contributing data / Data contributor agrees to respect the Procedures for contributing data to the POST-TB-IPD:*

1. Country programs, institutions, or individuals with data interested in contributing to the POST-TB-IPD should contact the Data custodian (Jonathon Campbell,), or any member of the POST-TB-IPD Oversight Committee. There will be a preliminary discussion with data contributors regarding the type and number of participants, information available, and assessed outcomes. If the Data Custodian believes that the data appears eligible for inclusion in the POST-TB-IPD, a data sharing agreement will be signed.
2. The data contributor will verify whether they require approval from their local Research Ethics Board, depending on their institution’s policy. If so, the data contributor will obtain this approval before sending the data to the Data Custodian (McGill Group). No additional data will be collected from the participants; thus, the data contributor will not need to obtain patients’ consent for this analysis.
3. Respect this data sharing agreement between the data contributor (individual or institution) and the Data Custodian.
4. Contact: the Data Contributor will designate one person who will be the primary point of contact with the Data Custodian. This is to ensure consistent and clear communication.
5. Instructions for data preparation: the Data Custodian will send to the data contributor detailed instructions, including a database template and data dictionary regarding how to format the data so they can most easily merge with the existing POST-TB-IPD. The data contributor will make every effort to prepare the data before transfer so that it meets these standards. If the Data contributor does not have sufficient technical capacity to format the data to the common standard, then the Data Custodian may undertake this - in specific instances and based on mutual agreement and when relevant resources are available.
6. Instructions for data transfer: the data contributor will send the data by secure file transfer, as mutually agreed upon with the Data Custodian to be NextCloud (<edit based on agreed platform>).
7. Data queries: when received by the Data Custodian, data will be checked for completeness and variable names and definitions. The data contributor will continue to work with the Data Custodian to resolve data problems. This may require significant work and time on the part of the data contributor.
8. Once the data is merged into the POST-TB-IPD, an update will be posted that reflects the date the data were added, the number of individual participant records, assessments performed (type and timing), and the WHO region of origin of the data contributing centre(s).
9. If the data contributed are used in an analysis, then attribution of the data source will be made in all public presentations or publications (at minimum an acknowledgement of the data contributor).
10. Consortium: The Data Contributor may designate themselves and at most one other person (i.e., a maximum of 2 people) to join “The Health and Wellbeing After Tuberculosis IPD Consortium.” This Consortium will be listed on all dissemination materials (e.g., presentations, publications).
11. Authorship: all members of the Consortium (defined above) will have the opportunity to actively participate and be recognized with authorship on analyses performed with their data. Being a member of the Consortium does not obligate authorship on publications and members are expected to meet all criteria for authorship as defined by the International Committee of Medical Journal Editors (ICMJE). Members of the Consortium may also nominate a maximum of one other individual to be considered for participation and authorship on each analysis. All authorship requests (whether by Consortium members or those they nominate) will be reviewed by the POST-TB-IPD Oversight Committee with a view for fairness and equity in authorship opportunities. Data Contributors who are not listed by name as authors will be captured within the Consortium name “The Health and Wellbeing After Tuberculosis IPD Consortium” on all publications, and included in the acknowledgments.
12. Data contributor will participate in the selection of members of the POST-TB-IPD Oversight Committee, comprised of representatives of data contributors and the Data Custodian. This committee will review proposed uses of the data (other than for WHO guidelines) including a detailed statistical analysis plan (SAP). The SAP will be reviewed by the POST-TB-IPD Oversight Committee who will approve, request revisions, or reject. A written response to each request must be made within 30 days.
13. Preliminary results: Data contributor may receive preliminary results of analyses using their data. All such results must be treated confidentially. The Investigator will not publish (including posting on the internet), present in any public forum, nor disseminate through any media these results without approval from the group performing the analyses, and the POST-TB-IPD Oversight Committee.

Dr. Jonathon Campbell (for the McGill University Group)	Date

*** Insert name and institution ***	Date

**ANNEX 1. POST-TB-IPD OVERSIGHT COMMITTEE**

Membership

The POST-TB-IPD Oversight Committee will encompass 7 members, plus the Data Custodian (Jonathon Campbell), who will only participate in votes in the event of an unresolvable decision. Initially, the 7 members will include the study principal investigators (Dr. Silvia Chiang, Dr. Marieke van der Zalm, and Dr. James Johnston) and 4 members selected by Data Contributors. The initial term of the membership will be 4 years and thereafter will be renewable on a 2-year basis. There is no remuneration for being a member of the Oversight Committee.

Selection Process

The only permanent member of the POST-TB-IPD Oversight Committee is the Data Custodian (Jonathon Campbell, who holds a generally non-voting role). The remaining members will be selected by majority vote among the Data Contributors. To be up for “selection,” a Data Contributor must volunteer to be considered to prevent individuals without time or interest being elected.

Roles

The POST-TB-IPD Oversight Committee will serve three specific roles:

1. Review requests for new individuals to have access to the POST-TB-IPD for the purposes of agreed upon analyses by the group.
2. Review and discuss proposals for new projects by Data Contributors or institutions/organizations, with final say on whether they are approved based on feasibility, novelty, and potential impact.
3. Review opportunities and requests for authorship and/or participation by Data Contributors on project proposals to ensure opportunity is equitable.
